# Molecular Profiling of the Hippocampus of Children with Autism Spectrum Disorder

**DOI:** 10.1101/2022.10.13.22281011

**Authors:** Lindsay Rexrode, Joshua Hartley, Kurt C Showmaker, Lavanya Challagundla, Michael W. Vandewege, Brigitte E. Martin, Estelle Blair, Ratna Bollavarapu, Rhenius B. Antonyraj, Keauna Hilton, Alex Gardiner, Jake Valeri, Barbara Gisabella, Michael Garrett, Theoharis C. Theoharides, Harry Pantazopoulos

## Abstract

Several lines of evidence point to a key role of the hippocampus in Autism Spectrum Disorders (ASD). Altered hippocampal volume and deficits in memory for person and emotion related stimuli have been reported, along with enhanced ability for declarative memories. Mouse models have demonstrated a critical role of the hippocampus in social memory dysfunction, associated with ASD, together with decreased synaptic plasticity. Chondroitin sulphate proteoglycans (CSPGs), a family of extracellular matrix molecules, represent a potential key link between neurodevelopment, synaptic plasticity, and immune system signaling. There is a lack of information regarding the molecular pathology of the hippocampus in ASD. We conducted RNAseq profiling on postmortem human brain samples containing the hippocampus from male children with ASD (n=7) and normal male children (3-14 yrs old), (n=6) from the NIH NeuroBioBank. Gene expression profiling analysis implicated molecular pathways involved in extracellular matrix organization, neurodevelopment, synaptic regulation, and immune system signaling. qRT-PCR and Western blotting were used to confirm several of the top markers identified. The CSPG protein BCAN was examined with multiplex immunofluorescence to analyze cell-type specific expression of BCAN and astrocyte morphology. We observed decreased expression of synaptic proteins PSD95 (p<0.02) and SYN1 (p<0.02), increased expression of the extracellular matrix (ECM) protease MMP9 (p<0.03), and decreased expression of MEF2C (p<0.03). We also observed increased BCAN expression with astrocytes in children with ASD, together with altered astrocyte morphology. Our results point to alterations in immune system signaling, glia cell differentiation, and synaptic signaling in the hippocampus of children with ASD, together with alterations in extracellular matrix molecules. Furthermore, our results demonstrate altered expression of genes implicated in genetic studies of ASD including SYN1 and MEF2C.

## Introduction

Autism Spectrum Disorder (ASD) is a developmental condition impacting the global population with increasing prevalence ^1^. ASD is characterized by impaired social interactions and communication along with stereotypic movements ^2, 3^. The pathogenesis of ASD is unknown, limiting development of therapeutic and preventative strategies. Several lines of evidence point to a key role of the hippocampus in ASD. Altered hippocampal volume and deficits in memory for person and emotion related stimuli have been reported, along with enhanced ability for declarative memories ^4, 5^. Specifically, increased hippocampal volume was reported in children and adults with ASD ^6^. Preclinical models have demonstrated a critical role of the hippocampus in social memory dysfunction associated with ASD, together with decreased synaptic plasticity ^7, 8^. Abnormalities in neurodevelopmental and neuroimmune processes are key features of ASD ^9, 10^ and may contribute to hippocampal dysfunction. Growing evidence indicates that altered neuroimmune signaling during development is critically involved in ASD ^11-16^. Brain imaging and human postmortem studies suggest that enhanced neuroimmune signaling is present in several brain regions in ASD ^12, 13, 16^. Furthermore, a recent single cell RNAseq profiling study of the prefrontal and anterior cingulate cortex implicates several microglial specific molecules ^17^. Maternal immune activation results in sex-specific microglial alterations primarily impacting male offspring ^14, 15, 18^, reflective of the male prevalence of ASD ^19^. Brain neuroimmune signaling has several distinctions from classic peripheral inflammatory processes that relate to aspects of ASD ^20^. Brain neuroimmune molecules are involved in a range of regulatory processes including synaptic plasticity and neurodevelopmental processes in addition to neuroimmune response ^20^, which may impact neurodevelopmental and synaptic regulation processes in ASD.

Chondroitin sulphate proteoglycans (CSPGs) and their endogenous proteases are critically involved in mediating immune responses and represent key factors at the intersection of neuroimmune signaling, neurodevelopment, and synaptic regulation. Chondroitin sulphate is a potent inhibitor of immune response (for review see ^21^). Several lines of evidence indicate that altered neurodevelopment plays a key role in ASD ^10, 22, 23^. For example, neuronal and dendritic spine development is altered in the amygdala of children with ASD ^9, 10^. Abnormalities in neuronal migration have also been reported ^22, 23^. CSPGs are extracellular matrix molecules (ECMs) critically in involved in neuronal migration, glial cell maturation, and synaptic stabilization ^24-26^. Work from our group and others have reported alterations ECMs including CSPGs in subjects with schizophrenia ^27, 28^. Genetic factors and gene expression pathways implicated in ASD share a degree of overlap with schizophrenia ^29, 30^. Recent evidence suggests these disorders may also share alterations in ECMs. Genome-wide association studies (GWAS) implicate several ECM genes in ASD, including genes encoding for endogenous proteases such as ADAMTS3, ADAMTS5, ADAMTS14 ^31-35^. Increased levels of the CSPG protease matrix metalloproteinase 9 (MMP9) have been reported in amniotic fluid samples of children with ASD ^36^.

Despite the critical role of CSPGs in neurodevelopment, neuroimmune signaling and synaptic regulation ^21, 37, 38^, and evidence for their involvement in several psychiatric disorders ^27, 39-41^, the role of CSPGs in the brain of children with ASD has not been examined. We propose that CSPGs are shared downstream targets from several genetic and environmental factors that are at the intersection of neurodevelopment, neuroimmune signaling, and synaptic regulation. As a first step in identifying the molecular pathology of the hippocampus in ASD, we conducted RNAseq profiling of human postmortem hippocampus samples from male children with ASD and age matched normotypic control subjects (3-14 yrs old) (Table 1). We focused on a neurodevelopmental time-window encompassing stages of synaptic development and refinement.

**Table 1:** Subjects and demographic information.

| Age | Sex | Race | RIN | PMI<br>(hrs) | Hemisphere | Epilepsy | Cause of<br>Death | Measures<br>(RNAseq+qPCR<br>, Western,<br>Microscopy) |
| --- | --- | --- | --- | --- | --- | --- | --- | --- |
| 5-10 | Male | Black | 7.4 | 18 | Left | No | natural | Western |
| 5-10 | Male | Black | 8.4 | 16 | Left | No | natural | RNA, Western,<br>Microscopy |
| 10-15 | Male | Black | 6.8 | 16 | Left | No | natural | RNA, Western,<br>Microscopy |
| 10-15 | Male | Black | 6.8 | 20 | Left | No | undetermined | RNA, Western,<br>Microscopy |
| 10-15 | Male | Hispanic | 2.4 | 15 | Left | No | not available | RNA, Western,<br>Microscopy |
| 10-15 | Male | White | 7.5 | 13 | Left | No | accidental | Western |
| 5-10 | Male | White | 6.9 | 12 | Left | No | accidental | RNA, Western,<br>Microscopy |
| 5-10 | Male | Black | 7.6 | 16 | Left | No | accidental | RNA, Western,<br>Microscopy |
| mean<br>± SD |  |  |  |  |  |  |  |  |
| 9.6±<br>3.25 | 8 M | 5B, 2W,<br>1H | 6.7±<br>1.8 | 15.8<br>±2.5 | 8 Left | 8 No |  |  |
| Age | Sex | Race | RIN | PMI<br>(hrs) | Hemisphere | Epilepsy | Cause of<br>Death | Measures<br>(RNAseq+qPCR<br>, Western,<br>Microscopy) |
| 5-10 | Male | N/A | 7.1 | 12 | Left | N/A | N/A | RNA, Western |
| 5-10 | Male | Hispanic | 7.1 | 27 | Left | No | natural | RNA, Western,<br>Microscopy |
| 5-10 | Male | Black | 6.7 | 20 | Left | No | accidental | Western |
| 5-10 | Male | White | 7.7 | 3 | Left | No | natural | RNA, Western,<br>Microscopy |
| 5-10 | Male | White | 6.0 | 21 | Left | No | accidental | RNA, Western,<br>Microscopy |
| 10-15 | Male | N/A | 4.8 | 20 | Left | N/A | N/A | RNA, Western,<br>Microscopy |
| 10-15 | Male | Black | 8.3 | 22 | Left | Yes | natural | RNA, Western,<br>Microscopy |
| 10-15 | Male | N/A | 6.5 | 15 | Left | N/A | N/A | RNA, Western,<br>Microscopy |
| mean<br>± SD |  |  |  |  |  |  |  |  |
| 8.6±<br>2.83 | 8 M | 2B, 2W,<br>1H, 3<br>N/A | 6.8±<br>1.1 | 17.5<br>±7.4 | 8 Left | 4 No, 1<br>Yes, 3<br>N/A |  |  |

## Methods

### Human Subjects

Postmortem human brain hippocampal samples of ASD (n=7) and non-ASD (n=6) male children (3-14 yrs old, Table 1) were obtained from the NIH NeuroBioBank at the University of Maryland, Baltimore MD which obtained informed consent. IRB approval was obtained from the Univ. of Maryland IRB committee. Cohort size was determined based on previous studies ^16^. Available samples for RNA analysis consisted of ASD (n=7) and non-ASD (n=6). For Western Blotting studies, hippocampal protein samples were available from the same subjects along with three additional samples, resulting in ASD (n=8) and non-ASD (n=8) male children (Table 1). Microscopy studies were conducted on a subset of frozen tissue sections from the same subjects consisting of 6 subjects with ASD and 6 control subjects in order to obtain cell type specific data and glial cell morphology information. Sections from the remaining subjects in the cohort did not display suitable cytoarchitecture for microscopy. Frozen blocks were sectioned at 30 µm thickness using a Leica CM 3050 S cryostat (Leica Biosystems, Buffalo Grove, IL). For fluorescent microscopy studies, sections were stained with DAPI to identify hippocampal subregions and to confirm cell specific labeling. This study is limited to males only because ASD is four times more common in males than females, and to avoid any additional sex and hormonal variabilities.

#### RNAseq Profiling

RNA isolation, library preparation, and next generation sequencing was performed by the Molecular and Genomics Core Facility at the University of Mississippi Medical Center, as described previously ^42^. Total RNA was isolated from tissue samples using the Invitrogen PureLink RNA Mini kit with Trizol (Life Technologies; Carlsbad, CA, USA) following manufacturer protocol. Quality control of total RNA was assessed using the Qiagen QIAxcel Advanced System for quality and Qubit Fluorometer for concentration measures. The RQI was 6.6 ± 2.1 (mean ± SD). Libraries were prepared using the TruSeq Stranded Total RNA LT Sample Prep Kit from Illumina (San Diego, CA, USA) per manufacturer’s protocol using up to 1 ug of RNA per sample. Libraries were index-tagged, pooled for multiplexing (all 13 samples) and sequencing was performed on the Illumina NextSeq 500 platform using a paired-end read (2 x75 bp) protocol with the Illumina 150 cycle High-Output reagent kit.

#### RNAseq Bioinformatics Analysis

Differential expression of genes (i.e. mRNA and non-coding RNA) was assessed between subjects with ASD and controls. Reads were aligned to the NCBI GRCh38Decoy Refseq genome with the basespace application RNA-Seq Alignment (Version: 2.0.1 [workflow version 3.19.1.12+master]) that conducted both splice aware genome alignment with STAR alignment (version 2.6.1a, ^43^) and transcriptome quantification with Salmon (version 0.11.2, ^44^). Differential expression (DE) of the genes (DGE) were conducted with the DESeq2 (1.20.0, ^45^) and tximport (version 1.8.0, Soneson 2015) R packages using the BaseSpace RNA-Seq Differential Expression (version 1.0.1 [Illumina Secondary Analysis Software version 3.18.18.9+master]) application. A gene was considered differentially expressed if the False Discovery Rate (FDR) adjusted p-value did not exceed 0.05. Heatmap visualizations were generated using the pheatmap (Kolde 2012) function in R based on hierarchical clustering (method=average; distance=correlation) of the ‘regularized log’ transformation values from DESeq2. Gene Set Enrichment Analysis (GSEA) was conducted with the ClusterProfiler R package ^46^ using the functions gseGO, gseKEGG, and gsePathway ^47^ for GO Biological Processes (BP), KEGG, and Reactome geneset collections, respectively, each with the parameters pAdjustMethod = “BH”, pvalueCutoff = 0.05, minGSSize = 20, and maxGSSize = 500. Plots were generated with R packages ggplot2 (Wickham 2016) and pheatmap (Kolde 2012). Raw sequencing reads were deposited into the NIH Data Archive Collection (C3917: experiment ID:2219 (DOI): 10.15154/1528650).

#### qRT-PCR

Total RNA (0.5 μg) was used as a template for synthesis of cDNA in a total reaction volume of 20 μl using Invitrogen iScript™ Advanced cDNA synthesis kits (cat# 1725038, Invitrogen, Grand Island, NY). Twenty-eight transcripts identified as differentially expressed in RNAseq studies were chosen for validation as well as the downstream protease MMP9 and the neuroimmune signaling molecule IL1B. B2M and ACTB were used as reference genes. Pre-validated qPCR probes from Bio-Rad PrimerPC were used (see Supplementary table 2). qPCR was performed using 384 well plates with the Bio-Rad CFX 384 real-time PCR detection system and iQ-SYBR Green Supermix (cat# 1708880, Bio-Rad, Hercules, CA). PCR reactions contained 10μl of the SYBR Green PCR mix, 0.04μl of 100μM forward and reverse primers, 1μl of cDNA, in a final volume of 20μl using nuclease free water. For all primer pairs, PCR cycling conditions were 50°C for 2 min and 95°C for 2 min, followed by 50 cycles of 95°C for 15 sec, 60°C for 15 sec and 72°C for 1 min. PCR product quantification was performed by the relative quantification method^48, 49^ and expressed as standardized arbitrary units ^48^.

#### Western blot analysis

Protein levels of MMP9, IL1β, PSD-95, SYN1, MEF2C, and DGCR6 were determined by Western blotting analysis. Brain tissues were homogenized using lysis radio-immuno precipitation (RIPA) buffer in the presence of a protease inhibitor cocktails (Sigma-Aldrich, St. Louis, MO), followed by sonication using a Polytron (Brinkmann Instruments, Westbury, NY). Total protein concentration was determined by bicinchoninic acid assay (Thermo Fisher Scientific, Waltham, MA) with bovine serum albumin (BSA) as standard. The total cellular protein (30 μg aliquots) was separated using Biorad 4-15 % MP TGX Stain-Free gels under SDS denaturing conditions (Biorad, Hercules, CA) and electrotransfered onto Biorad LF PVDF membranes (Biorad, Hercules, CA) using Biorad rapid transfer kits. Blocking was carried out using Biorad EveryBlot blocking buffer with 0.01 % Tween-20 (Biorad, Hercules, CA). The membranes were probed with the following primary antibodies: rabbit anti-MMP9 (1:1000 µl, cat#AB13458, MilliporeSigma, Burlington, MA), rabbit anti-PSD-95 (1:500 µl, cat#20665-1-AP, Protein Tech, Rosemont, IL), rabbit anti-SYN1 (1:500 µl, cat#20258-1-AP, Protein Tech, Rosemont, IL), rabbit anti-MEF2C (1:500 µl, cat#10056-1-AP, Abnova, Taipei City, Taiwan), DGCR6 (1:500 µl, cat#H00008214-B01P, Protein Tech, Rosemont, IL) using VCP for the loading control (mouse anti-VCP, Santa Cruz Biotechnology cat# sc-57492 and rabbit anti-VCP, Abcam Inc. cat# ab111740) according to prior studies ^50^. All proteins were visualized with Biorad anti-mouse Starbright blue 520 and anti-rabbit Starbright blue 700 fluorescence secondary antibodies (Biorad, Hercules, CA). Blots were imaged on a Biorad ChemicDoc MP Imaging system and analyzed using Biorad Image Lab v 6.0.1 (Biorad, Hercules, CA).

#### Immunohistochemistry

Triple immunofluorescence labeling was performed using primary antibodies for the target proteins rabbit anti-MMP9 (1:500 µl, cat#AB13458, MilliporeSigma, Burlington, MA), rabbit anti-BCAN (1:500 µl, cat#19017-1-AP, Protein Tech, Rosemont, IL) biotinylated Wisteria floribunda agglutinin lectin (1:1000 µl, cat# B-1355, Vector Labs, Burlingame, CA), mouse anti-IBA1 (1:1000 µl, Wako Chemicals, cat# 013-27593), rabbit anti-IBA1 (1:1000 µl, Wako Chemicals cat#019-19741), or mouse anti-TPSAB1 (1:500 µl, cat#66174-1-Ig, Protein Tech, Rosemont, IL). Sections were post-fixed in 4% PFA for 30 minutes, and then co-incubated in primary antibodies in 2% bovine serum albumin (BSA) for 72 hr at 4 °C. This step was followed by 4 hour incubation at room temperature in Alexa Fluor goat anti-mouse 647 (1:300 µl; A-21235, Invitrogen, Grand Island, NY) and donkey anti-rabbit 555 (1:300 µl; A-32794, Invitrogen, Grand Island, NY), followed by 2 minutes in TrueBlack solution for autofluorescence quenching (Biotum, cat#23007) ^51, 52^. Sections were mounted and coverslipped using Dako mounting media (S3023, Dako, North America, Carpinteria, CA). All sections were coverslipped and coded for quantitative analysis blinded to diagnosis. Sections from all brains included in the study were processed simultaneously to avoid procedural differences. Omission of streptavidin or omission of the primary or secondary antibodies were used as negative controls.

#### Microscopy Quantification

Sections containing the hippocampus were quantified using an Olympus BX 61 fluorescent microscope interfaced with StereoInvestigator v11 (MBF Biosciences, Williston, VT). Borders of each subregion were defined according to the Allen Brain Atlas and traced under 4x magnification. Each traced region was systematically scanned through the full x, y, and z-axes under 40x magnification to count each immunolabeled cell.

#### Sholl Analysis

Fluorescent multichannel images were systematically sampled from CA4 of glial fibrillary acidic protein (GFAP) labeled glial cells with or without BCAN expression and captured using a 60x oil objective (369 glial cells from subjects with ASD and 419 glial cells from control subjects obtained from 6 ASD subjects and 6 control subjects) using StereoInvestigator v 11.0 (MBF Biosciences, Williston, VT). Images were analyzed using the Sholl analysis probe in Neurolucida 360 (MBF Biosciences, Williston, VT).

Numerical Densities of Immunoreactive Cells: Numerical densities were calculated as Nd= ∑N / ∑V where N = sum of all cells counted in each region, and V is the volume of each region, calculated as V= ∑a • z, where z is the thickness of each section (30 μm) and a is area in μm2.

#### Statistical Analysis

Stepwise linear regression analysis of covariance was applied to the main outcome qRT-PCR, Western blotting, and microscopy measures to test differences between ASD and control groups and effects of confounding variables including age, PMI, RIN, race, history of epilepsy, and cause of death. Logarithmic transformation was applied to values when not normally distributed. Potential effects of these confounding covariates were tested on our outcome measures in stepwise linear regression analysis, as conducted previously ^56^ using JMP Pro v15.1.0 software (Cary, North Carolina). Significance of comparisons by ANOVA is denoted by p<0.05. No information was available on severity of ASD, any comorbidities or the effects of any therapeutic drugs in the ASD group.

## Results

### RNAseq Profiling

RNAseq gene expression profiling identified 2851 differentially expressed genes in the hippocampus of children with ASD, including genes implicated in genetic analysis studies ^53-56^ and genes involved in synaptic regulation, blood vessel and blood-brain barrier regulation, immune signaling, ECM organization, and calcium channel activity (Fig. 1, Supplementary Fig.1, and Supplementary Table 1).

**Figure 1:**
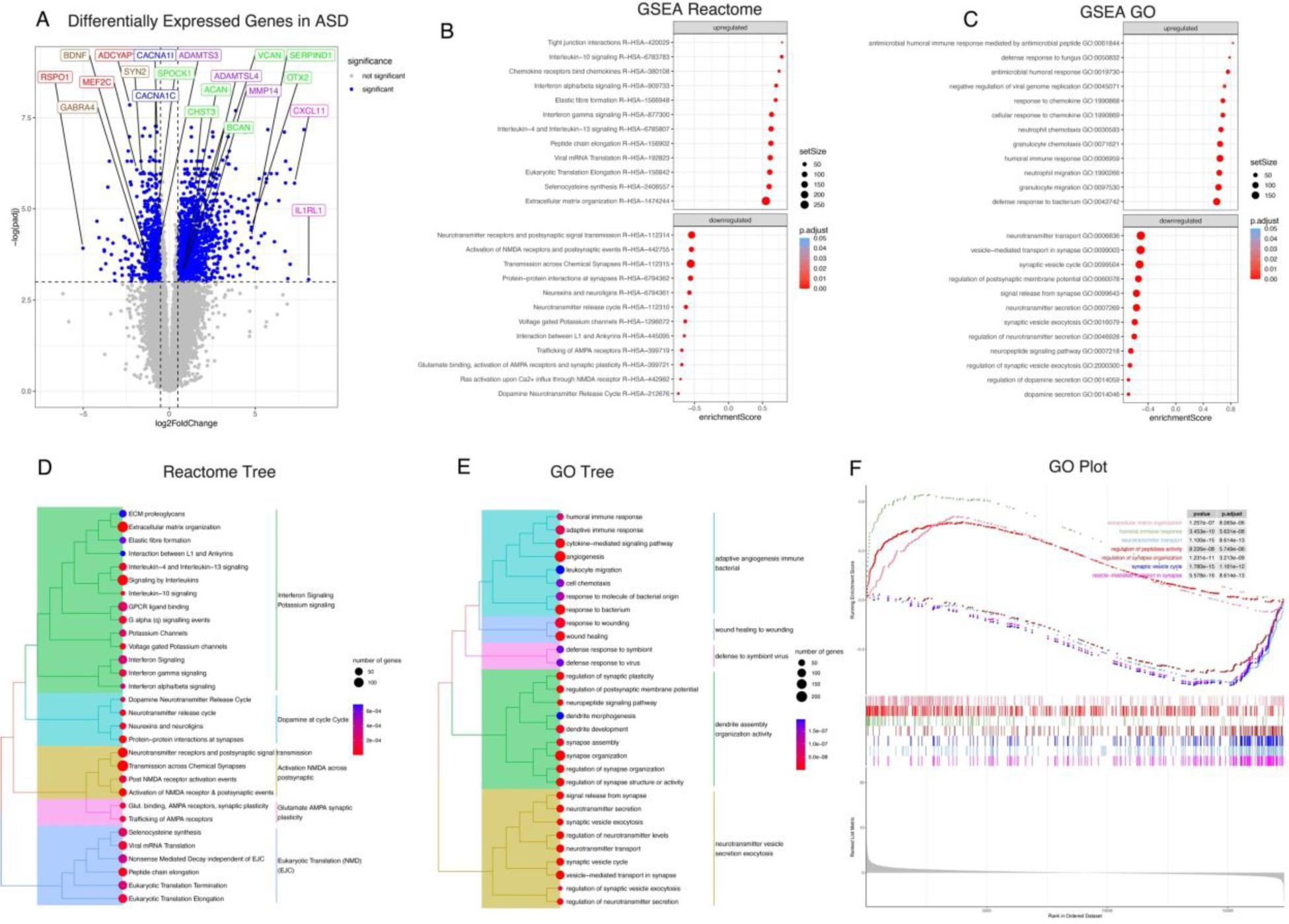
Differential Gene Expression Pathways in the Hippocampus of Children with ASD. (A) Volcano plot of DGE: ASD vs. non-ASD controls. On the x-axis is log fold change of genes in subjects with ASD compared to controls, points to the right of 0 represent genes that are increased, and points to the left of 0 genes that are decreased, in ASD compared to controls. Statistical significance is displayed on the y-axis, and p < 0.05 are labeled in blue. Select genes with both high fold change and significance are labeled. The top dozen GSEA Reactome analysis identified upregulated pathways, (including extracellular matrix organization and immune signaling pathways), and downregulated pathways involved in synaptic regulation in children with ASD (B). Similar top pathways were detected with GSEA GO BP analysis (C). Reactome tree and GO tree plots of pathways altered in children with ASD (D&E). GSEA GO enrichment plot of upregulated and downregulated pathways in children with ASD (F).

Pathway analysis identified upregulation of pathways involved in immune and inflammatory response (ex:R-HSA-877300; GO0006959), vasculature regulation (ex:hsa04610) and ECM organization (ex: R-HSA-1474244) (Fig. 1, Supplementary Fig.1, and Supplementary Table 1). We identified downregulation of several pathways involved in synaptic signaling and synaptic transmission (ex:GO0099504; R-HSA-112314) (Fig. 1, Supplementary Figs.1&2, and Supplementary Table 1). Hierarchical cluster analysis revealed expression variability among subjects and DEGs (Fig. 2 A). However, there was noticeable separation between control and ASD subjects among ECMs, ECM proteases, and synaptic signaling molecule genes, but weaker clustering among genes related to immune signaling (Fig. 2 B-E).

**Figure 2:**
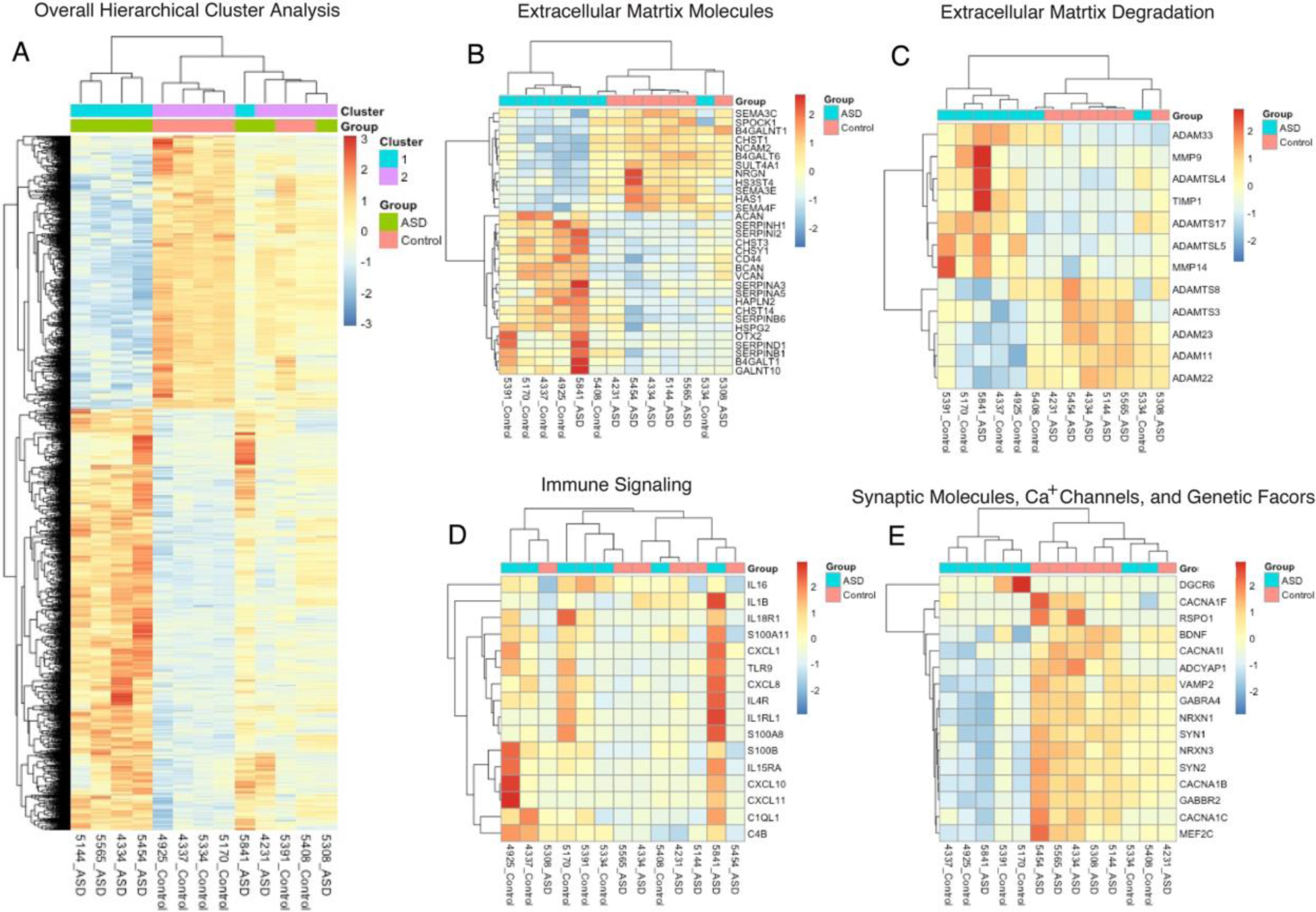
Hierarchical Cluster Analysis. (A) Hierarchical and K means clustering of 7 ASD and 6 non-ASD control subjects using regularized log expression of 2851 genes differentially expressed between ASD and non-ASD control subjects in the hippocampus (p-adjusted < 0.05). Hierarchical clustering of differentially expressed genes for extracellular matrix molecules (B), extracellular matrix degradation molecules (C), immune signaling (D), and synaptic molecules and genetic factors (E).

### qRT-PCR Confirmation of DEGs

qRT-PCR on a subset of differentially expressed genes (DEGs) confirmed several of the top DEGs. Genes highly expressed in the hippocampus of children with ASD included IL1RL1, MMP9, OTX2 and SERPIND1 (Fig. 3A). Increased expression was also detected for several additional ECMs including ADAMTSL4, CHST3, MMP14 and VCAN, and decreased expression for ADAM23, ADAMTS3, SEMA3E, and SPOCK1 (Fig. 3B). Decreased expression was observed for CACNA1I, whereas expression for additional calcium channels and IL1beta approached significance (Fig. 3C). Decreased expression was confirmed for the genetically associated markers SYN1 and ADCYAP1 (Fig. 3D), whereas DGRC6 and RSPO1 did not approach significance between groups. Decreased expression was confirmed for most of the synaptic markers examined (Fig. 3E).

**Figure 3:**
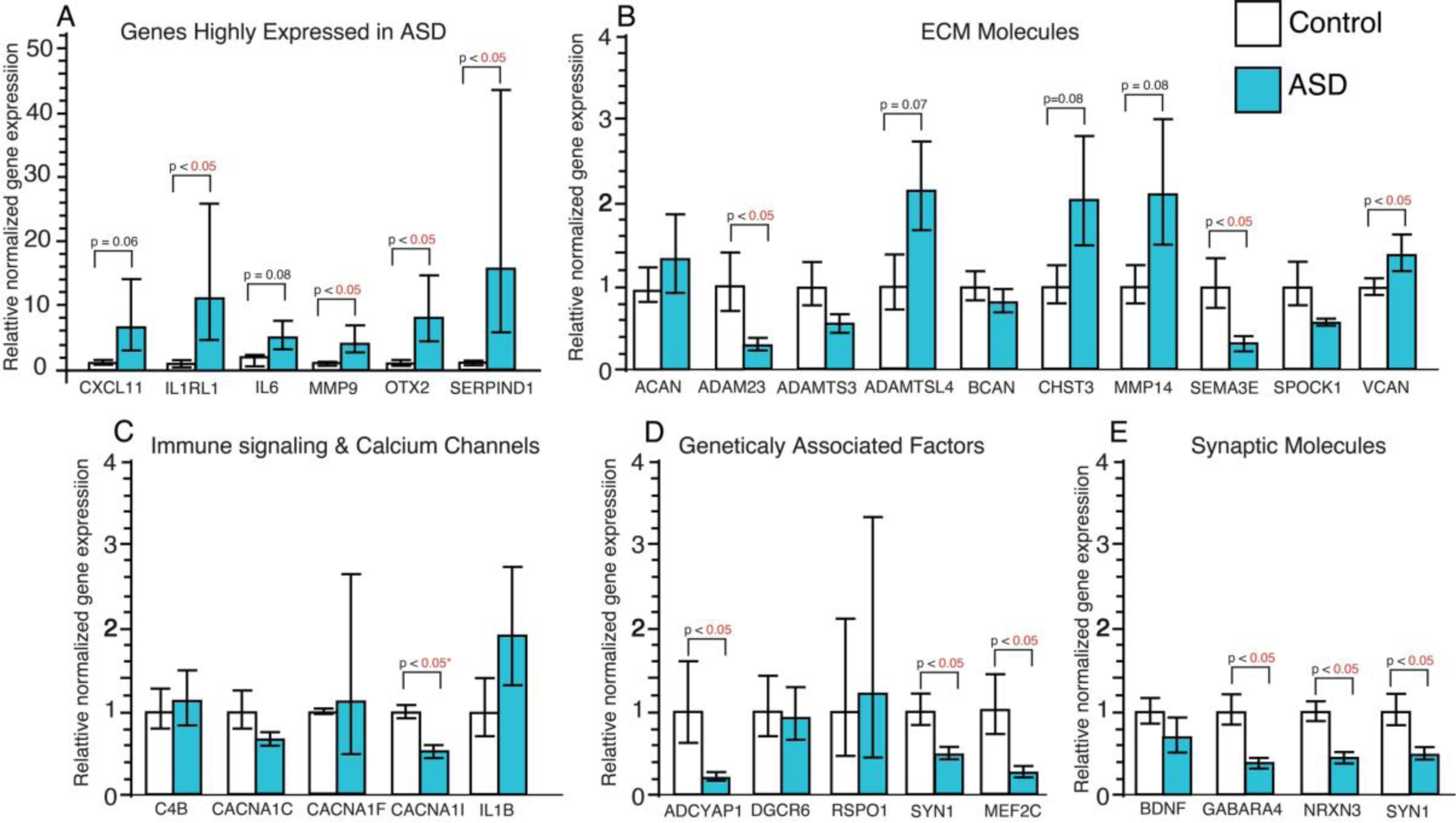
qPCR Confirmation of Differentially Expressed Genes. qPCR analysis was conducted on a subset of 28 genes with differential expression from our RNAseq analysis as well as the downstream extracellular matrix protease MMP9 and the neuroimmune signaling gene IL1B. Significantly increased expression was detected for genes highly expressed in the hippocampus of children with ASD (A). Altered expression was confirmed for several extracellular matrix molecules (B) and the L-type Ca channel CACNA1I (C). Altered gene expression was also confirmed for several genes implicated as genetic factors for ASD (D) and for synaptic molecules (E). *adjusted for significant effects of age and PMI. Significance values are derived from stepwise linear regression models. Bar graphs depict the mean (histogram) and 95% confidence intervals (black lines).

### Western Blotting Protein Confirmation of DEGs

Western blotting on several of the top candidates confirmed gene expression measures with protein levels. The active and cleaved forms of MMP9 were increased in children with ASD whereas the precursor form was not significantly different between the two groups (Fig. 4 A-C). The 17 and 30 kDa isoforms of IL1beta protein were both significantly greater in children with ASD compared to age matched control subjects (Fig. 4 D&E). Furthermore, decreases in the synaptic proteins PSD95 and SYN1 were observed in subjects with ASD (Fig. 4 F-H). A significant decrease in protein expression was observed for MEF2C (Fig. 4I) but not for DGRC6 (Fig 4J).

**Figure 4:**
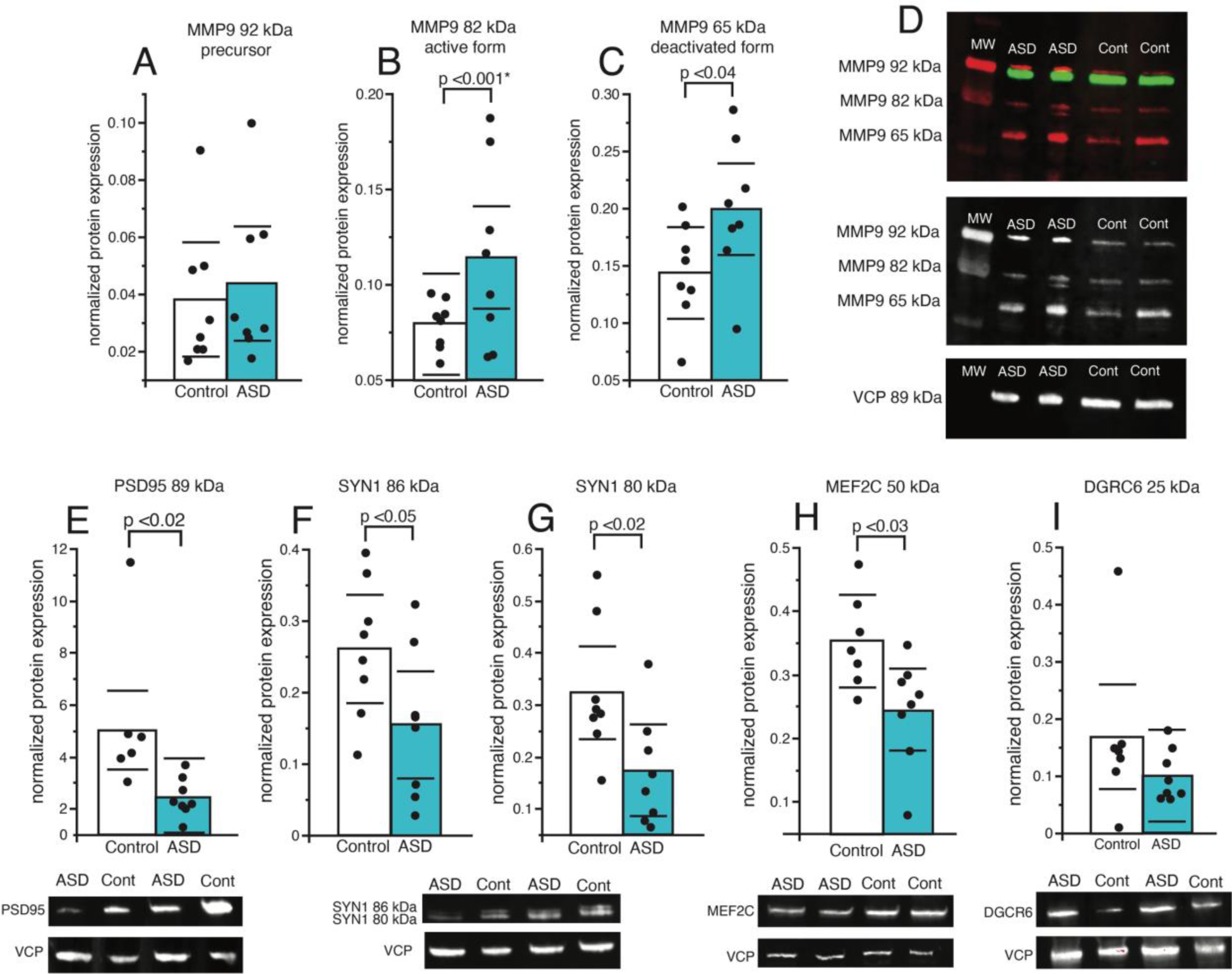
Protein Analysis of MMP9, Synaptic Markers and Genetically Associated Molecules. Western blot analysis was conducted on the extracellular matrix protease MMP9 and the synaptic markers PSD95 and SYN1, as well as the genetically associated molecules MEF2C and DGCR6. (A) No difference was observed for the 92 kDa isoform of MMP9, whereas expression of the 82 kDa (B) and 65 kDa (C) isoforms were increased in children with ASD. (D) Representative Western blots of MMP9. Expression of the broad postsynaptic marker PSD95 was decreased in children with ASD (E), along with decreased expression of the 86 kDa (F) and 80 kDa (G) isoforms of the synaptic marker SYN1. Expression of MEF2C, a molecule associated with ASD in genetic studies, was significantly decreased in children with ASD (H), whereas no difference was observed for expression of genetically associated molecule DGCR6 (I). Significance values are derived from stepwise linear regression models. Bar graphs depict the mean (histogram), black circles depict values for each subject and black lines the 95% confidence intervals.

### MMP9 Expression in Microglia, Neurons and Mast Cells

MMP9 labeling in the hippocampus was observed primarily in microglia a well as a small percentage of neurons and cells displaying mast cell morphology. Tryptase alpha/beta1 (TPSAB1) labeling was used to confirm the presence of mast cells in the human hippocampus of subjects with ASD and control subjects (Supplementary Fig. 3 A-C). Immunofluorescence confirmed that MMP9 expression colocalized with mast cells (Supplementary Fig. 3 D) and microglial cells labeled with IBA1 (Supplementary Fig. 3E). Densities of MMP9 labeled mast cells were not altered in the DG or CA4 of subjects with ASD compared to control subjects (Supplementary Fig. 3 F-H).

### Increased BCAN Expression in GFAP+ Astrocytes in Children With ASD

Brevican (BCAN) expression was observed in cells with astrocytic morphology, and 20-30% of these GFAP cells displayed co-expression of BCAN (Supplementary Fig. 4 A&B). Quantification of BCAN+ and GFAP+ cells in the CA4 area of the hippocampus revealed increase density of GFAP cells co-expressing BCAN (Supplementary Fig. 4C). Densities of cells expressing BCAN only or GFAP only were not altered between groups, suggesting that a greater percentage of GFAP cells express BCAN in children with ASD. Statistical comparison of the percentages of GFAP cells expressing BCAN between groups provides additional support for increased expression of BCAN in GFAP cells in children with ASD (Supplementary Fig. 4F).

### Altered Astrocyte Morphology in the Hippocampus of Children with ASD

Sholl analysis was performed on images systematically sampled from CA4 of GFAP labeled glial cells with or without BCAN expression (369 glial cells from subjects with ASD and 419 glial cells from control subjects obtained from 6 subjects per group) in order to test the hypothesis that astrocytes expressing BCAN display morphological features characteristic of immature glial cells. Glial cells were traced using the Sholl analysis probe in Neurolucida 360 (Supplementary Fig. 5 A&B) and branches were quantified for measures of branch intersections, length, surface area, volume, diameter, nodes and endings (Supplementary Fig. 5C). In control subjects, GFAP astrocytes co-expressing BCAN had significantly fewer branch intersections along with decreased branch length, surface area, volume and diameter (Supplementary Fig. 5 D-J). In comparison, GFAP astrocytes co-expressing BCAN from subjects with ASD had fewer branch intersections, nodes, and endings, along with decreased branch length (Supplementary Fig. 5 K-P). Direct comparison between ASD and control subjects revealed increased branch nodes and endings in GFAP only astrocytes in children with ASD compared to decreased branch nodes and endings in GFAP astrocytes co-expressing BCAN (Fig.4). A significant interaction between age and diagnosis was observed for branch intersections across all astrocytes examined, resulting in an opposite correlation of age with branch intersections in children with ASD compared to non-ASD control subjects (Fig. 4Q).

## Discussion

Our results represent, to our knowledge, the first evidence for molecular abnormalities in the hippocampus of children with ASD. We detected gene expression differences between ASD and control subjects and many changes were linked to ECM regulation, neuroimmune signaling, and decreased synaptic signaling are in line with growing evidence for neuroimmune signaling ^11-16^ and synaptic pathology in ASD ^10, 17, 54, 57, 58^. Our results highlight the potential involvement of ECMs with these pathways during a window of neurodevelopment in the hippocampus of children with ASD. Cluster analysis displayed the expected variability in subjects with ASD but suggests that ECMs and synaptic signaling molecules encompass a larger group of subjects than neuroimmune molecules. Furthermore, our data suggest that altered expression of the ECM BCAN may be associated with glial cell maturation deficits in ASD. Several genes implicated by genetic studies on ASD, including MEF2C and SYN1 ^53, 54, 58, 59^, displayed altered expression in our study, suggesting that these genetic factors may in part contribute to molecular pathology in the hippocampus of children with ASD.

### Extracellular Matrix Molecules

Gene pathways involved in ECM organization were upregulated in the hippocampus of children with ASD (Fig. 1; Reactome pathway), together with mRNA and protein expression changes confirmed with qRT-PCR and Western Blotting for several ECMs (Figs. 3&4). CSPGs and their endogenous proteases are critically involved in mediating neurodevelopment, synaptic regulation, and neuroimmune signaling, and thus represent key factors at the intersection of these processes in ASD.

Several ECMs upregulated in children with ASD may contribute to neuroimmune signaling processes. Chondroitin sulphate (CS) is a potent inhibitor of immune response ^21, 38^ and also inhibits human mast cells ^60^, activation of which has been implicated in ASD ^61^. CS protects from inflammatory neurodegeneration and promotes CNS repair ^62, 63^. Genetic reduction of chondroitin sulphate synthase 1 (CHSY1) causes neuroinflammation and neurodegeneration in the mouse hippocampus ^38^. Increased expression of ECM proteases may also contribute to enhanced neuroimmune signaling and blood-brain barrier permeability. The endogenous CSPG proteases matrix metalloproteinases (MMPs) are critically involved in promoting neuroimmune signaling ^64, 65^. Increased levels of the CSPG protease matrix metalloproteinase 9 (MMP9) have been reported in amniotic fluid samples of children with ASD ^36^. MMPs, including MMP9, are primarily produced by astrocytes and microglia in the brain ^66, 67^. MMP9 is also produced by mast cells ^68, 69^, and our data shows MMP9 expression predominantly in microglia and to a lesser extent in neurons and mast cells in the hippocampus of children with ASD (Supplementary Fig. 2).

Evidence from animal models supports the hypothesis that increased MMP9 expression during development contributes to decreased perineuronal nets (PNNs) and synaptic destabilization ^70-72^. PNNs are ECM structures that develop around fast-firing neurons and stabilize synaptic plasticity ^70-73^. Fragile-X syndrome is a monogenetic disease in which approximately 30% of patients display symptoms of ASD. Several animal models of Fragile-X syndrome show increased MMP9 expression and reductions of PNNs in the amygdala, auditory cortex and hippocampus, together with impaired fear memory ^70-73^. Pharmacological inhibition of MMP9 during development or genetic reduction of MMP9 in these mice restores PNN levels and reduces anxiety ^70, 71^, supporting the hypothesis that elevated MMP9 during development reduces CSPGs, impairs PNN development and in turn destabilizes synapses during this developmental window.

Altered expression of ECMs may also contribute to neurodevelopmental dysfunction in ASD. For example, VCAN, which encodes the core CSPG protein versican, promotes synaptic maturation during development ^74^, as well as neuronal differentiation and neurite outgrowth ^75^. CHST3, which was also upregulated in ASD, encodes the enzyme involved in chondroitin 6 sulphation and promotes hippocampal synaptic plasticity and memory ^76^. Our observed decreased expression of the chondroitin-heparan sulphate proteoglycan SPOCK1 in the hippocampus of children with ASD may contribute to neurodevelopmental dysfunction. SPOCK is highly expressed during brain development in areas of neuronal migration and axonal outgrowth, as well as in synaptic fields ^77^. We previously observed decreased SPOCK1 and SPOCK3 mRNA expression in the brain of subjects with schizophrenia ^78^, which shares genetic overlap with ASD ^79^. Furthermore, decreased SPOCK expression was correlated with decreased cognitive function in subjects with schizophrenia ^78^, suggesting that decreased SPOCK expression may be associated with cognitive function in ASD.

Increased expression of CSPGs may be a compensatory effect to increases in ECM proteases. Increased expression of CSPGs may also reflect their roles in neurodevelopmental processes such as cell maturation, as suggested by our data on BCAN expression in astrocytes. For example, BCAN is expressed in rat hippocampal astrocytes as they mature ^80^, suggesting BCAN expression promotes astrocyte development. Our observed increase of BCAN positive astrocytes in children with ASD together with impaired astrocyte morphology suggests that increased expression of CSPGs may be associated with immature glial cells in the hippocampus of children with ASD (Fig. 5, Supplementary Figs. 4&5).

**Figure 5:**
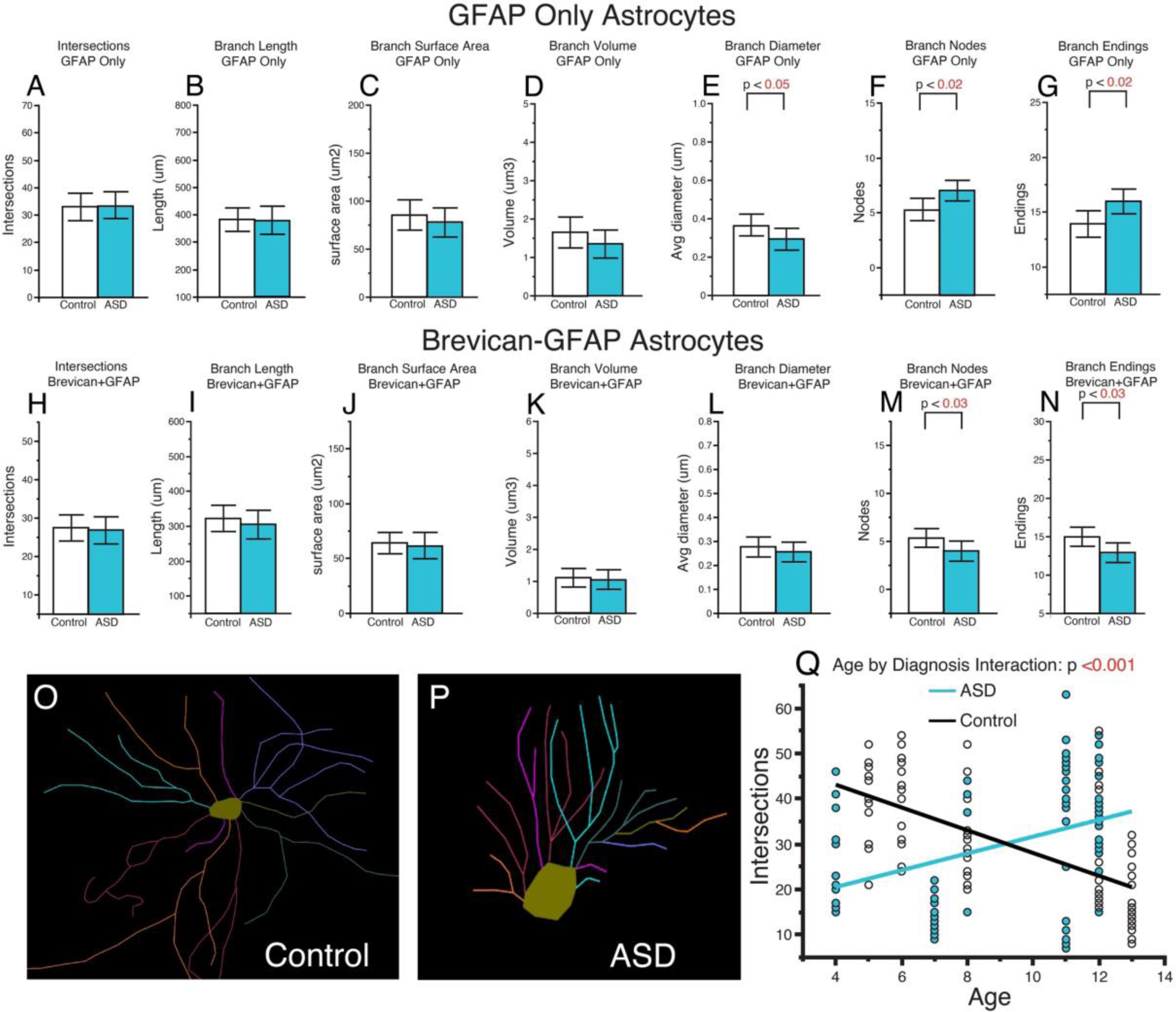
Altered Astrocyte Morphology in the Hippocampus of Children with ASD. Sholl analysis was conducted on a subset of samples from children with ASD and non-ASD control subjects (n=6/group). (A) No differences in branch intersections, branch length, surface area, or volume, were observed between diagnosis groups for astrocytes expressing GFAP only (A-D). A significant decrease was observed for branch diameter (E) together with an increase in branch nodes and endings (F&G) for GFAP only astrocytes in children with ASD. No differences were detected for branch intersections, branch length, surface area, volume, or diameter for astrocytes co-labeled with GFAP and BCAN (H-L). Branch nodes and endings were significantly decreases in GFAP-BCAN astrocytes from children with ASD (M&N). Representative tracings of a GFAP-BCAN astrocyte from a control subject (O) and a subject with ASD (P). Branch intersections across all astrocytes displayed a significant interaction between age and diagnosis, resulting in a negative correlation of intersections by age in control subjects compared to a positive correlation in subjects with ASD (Q). Blue circles represent values from subjects with ASD, white circles represent values from control subjects. Significance values are derived from stepwise linear regression models. Bar graphs depict the mean (histogram) and 95% confidence intervals (black lines).

### Synaptic Signaling

Our observed decreased gene expression in several pathways involved in synaptic regulation (Fig.1 and Supplementary Figures 1&2), as well as decreased mRNA and protein expression of synaptic markers together indicate decreased synaptic signaling in the developing hippocampus of children with ASD. We observed decreased protein expression of the synaptic marker PSD-95 (Fig. 4), which has been associated with NMDA receptor alterations and spine changes in ASD ^57^. Alterations in synaptic proteins suggest that synaptic alterations in the developing brain of children with ASD may arise from genetic factors such as reported genetic mutations for SYN1 associated with ASD ^54^. Evidence that loss of function genetic mutations in SYN1 have been associated with ASD and epilepsy ^54^, and SYN1 knockout mice display impaired social behaviors and repetitive behaviors, ^58^ supports this possibility. In addition, the observed increase of MMP9 expression and decreased expression of the synaptic markers PSD95 and SYN1 in the same cohort (Fig. 4), suggests that increased MMP9 in the developing brain of children with ASD may contribute to reductions of CSPGs that are involved in stabilizing synapses.

### Molecules Implicated as Genetic Factors in ASD

Several molecules implicated as genetic factors for ASD, including MEF2C, SYN1, DGCR6, displayed significantly altered gene expression in our RNAseq analysis. MEF2C and SYN1 decreased expression was also confirmed with qRT-PCR and Western blotting. MEF2C haploinsufficiency has been associated with ASD as well as epilepsy and intellectual disability ^53, 59, 81, 82^. Furthermore, mouse models of MEF2C haploinsufficiency demonstrate that impaired MEF2C function results in social deficits, reduced ultrasonic vocalization, hyperactivity, repetitive behavior, and synaptic regulation through both neuronal and microglial cells ^53, 83^. Our findings for decreased gene and protein expression of MEF2C in the hippocampus of children with ASD provide the first evidence for altered MEF2C expression in this region in ASD and support the involvement of MEF2C in this disorder. These results also suggest that MEF2C haploinsufficiency may contribute in part to several of the altered molecular pathways observed in our study. Genetic mutations in SYN1 resulting in loss of function have also been associated with ASD and epilepsy ^54^. Our observed decrease in SYN1 mRNA and protein expression suggests that SYN1 mutations may in part contribute to decreased SYN1 expression in the hippocampus of children with ASD.

DGCR6 has also been implicated as a genetic factor in ASD as part of the 22.q.11.22 deletion syndrome ^55^ and was one of the top DEGs in our RNAseq analysis (Supplementary Table 1). However, qRT-PCR and Western blotting measures did not detect changes in DGCR6 protein expression in the hippocampus of children with ASD (Figs 3&4). This may be due in part to the fact that RNAseq evaluates data across the entire transcript/gene and qRT-PCR only evaluates a single region of the gene that may not take into account possible splice variants. Genome-wide association studies (GWAS) implicate several ECM genes in ASD, including genes encoding for endogenous proteases such as ADAMTS3, ADAMTS5, ADAMTS14 ^32-35^, suggesting that genetic factors in ECMs may contribute to the broad ECM gene expression changes we observed in children with ASD.

### Neuroimmune Signaling

Neuroimmune molecules in the brain are key mediators of regulatory processes including synaptic plasticity and neurodevelopmental processes ^20^. Our findings of altered hippocampal neuroimmune signaling pathways may represent aspects of synaptic alterations and neurodevelopmental processes disrupted in children with ASD. These findings are in line with several studies suggesting a critical role for neuroimmune signaling during development in ASD, potentially contributing to synaptic abnormalities ^10, 11, 13, 14, 16, 54^. Increased cytokine levels during early developmental stages are associated with increased risk of developing ASD ^84, 85^. Rodent models of ASD suggest that maternal immune activation and early postnatal neuroimmune signaling contribute to synaptic dysfunction in several brain regions ^15, 86^, including the hippocampus ^87, 88^. Our findings for increased gene expression in inflammatory signaling pathways provide support for the involvement of neuroimmune signaling in the developing hippocampus of children with ASD. Several genes with significantly increased expression in our study have recently been implicated in astrocyte response to inflammation, including CXCL10, GBP2, TIMP1, SPERPINA3 ^89^. Several of these molecules including CXCL10, TIMP1 and well as IL1RL1 have been demonstrated to impact synaptic plasticity ^90-93^. ECMs are intricately involved with neuroimmune signaling in the regulation of neurodevelopmental processes and synaptic plasticity. A recent single cell RNAseq profiling study in postmortem samples of subjects with ASD implicated microglial alterations in ASD ^17^. These changes consisted of altered expression of ECMs involved in neurodevelopmental and synaptic regulation, including increased expression of the CSPG sulfotransferase CSGALNACT1 in microglia, and increased expression of the endogenous CSPG proteases MMP16 and ADAMTS9 ^17^. Furthermore, microglial signaling through the IL1RL1 receptor regulates hippocamal synaptic plasticity through ECM remodeling ^93^, providing further support tha neurommune signaling molecules altered in children with ASD may impact ECM and synaptic molecules. Altered neuroimmune signaling thus may be at the intersection of immune signaling, synaptic plasticity and neurodevelopmental processes in the hippocampus of children with ASD.

### Blood-Brain Barrier Regulation

Gene pathways involved in regulation of blood vessels were upregulated in children with ASD in our study (Fig. 1: angiogenesis GO pathway; Supplemental Fig. 1 and Supplemental Table 1). Furthermore, ECMs are also highly involved in blood-brain barrier (BBB) regulation ^94^. Our findings suggesting altered BBB composition in the hippocampus of children with ASD are in line with recent evidence for BBB dysfunction in ASD ^95, 96^. Altered expression of genes involved in BBB integrity were reported in postmortem cortex and cerebellum samples from subjects with ASD, together with increased expression of MMP9 and neuroimmune signaling molecules ^96^. Mutations in CHD7 have been associated with ASD and may contribute to changes in BBB glial cells that impact serotonin signaling and sleep defects ^95^. CHD7 gene expression was significantly upregulated in our RNAseq dataset (Supplementary Table 1). SERPIND1, upregulated in our dataset in children with ASD, is involved in promoting vascular endothelial function and angiogenesis ^97^, and is activated by glycosaminoglycans ^98^. Furthermore, a recent report demonstrated that decreased SPOCK1 expression results in increased BBB leakage in the developing mouse brain ^99^. We observed decreased SPOCK1 mRNA in hippocampal samples from children with ASD (Figs.1&3), suggesting that impaired SPOCK expression may contribute to BBB dysfunction in this disorder. Speculatively, our observed changes in ECM signaling pathways may contribute to altered BBB permeability and in turn increased neuroimmune signaling in the hippocampus of children with ASD.

### Technical Considerations

Our study consisted of bulk hippocampal gene and protein expression analyses, which does not allow for analysis of expression changes in specific hippocampal subfields or anterior to posterior gradients in expression. Furthermore, our bulk hippocampal profiling approach did not allow for evaluation of cell type specific changes. Future studies consisting of hippocampal subregion specific profiling and larger numbers of subjects may provide greater information regarding the hippocampal neurocircuitry alterations in children with ASD.

In summary, our results provide evidence regarding molecular alterations in the hippocampus of children with ASD during a key neurodevelopmental period. These findings point to ECM abnormalities at the intersection of gene expression pathways involved in synaptic regulation, blood-brain barrier regulation, and neuroimmune signaling. Several factors implicated in genetic studies of ASD including MEF2C and SYN1 displayed altered gene expression in our study and suggest that alterations in signaling pathways involved in ECM, neuroimmune signaling and synaptic regulation may be downstream from multiple genetic factors in ASD.

## Acknowledgments

We thank the NIH NeuroBioBank (https://neurobiobank.nih.gov/) for making the human brain samples available. The authors deeply appreciate the invaluable contributions made by the families consenting to donate brain tissue. The work performed through the UMMC Molecular and Genomics Facility is supported, in part, by funds from the NIGMS, including the Molecular Center of Health and Disease (P20GM144041), Mississippi INBRE (P20GM103476) and Obesity, Cardiorenal and Metabolic Diseases-COBRE(P30GM149404). Funding for these studies was also provided by the Inflammation Healing Foundation.

## Data Availability Statement

Gene expression profiling data will be publicly available on NCBI dbGaP upon manuscript publication. All other data are available in the main text or supplementary materials.

