## Supplemental Materials for "Molecular Profiling of the Hippocampus of Children with Autism Spectrum Disorder"

Rexrode et al Supplemental Figures and Legends

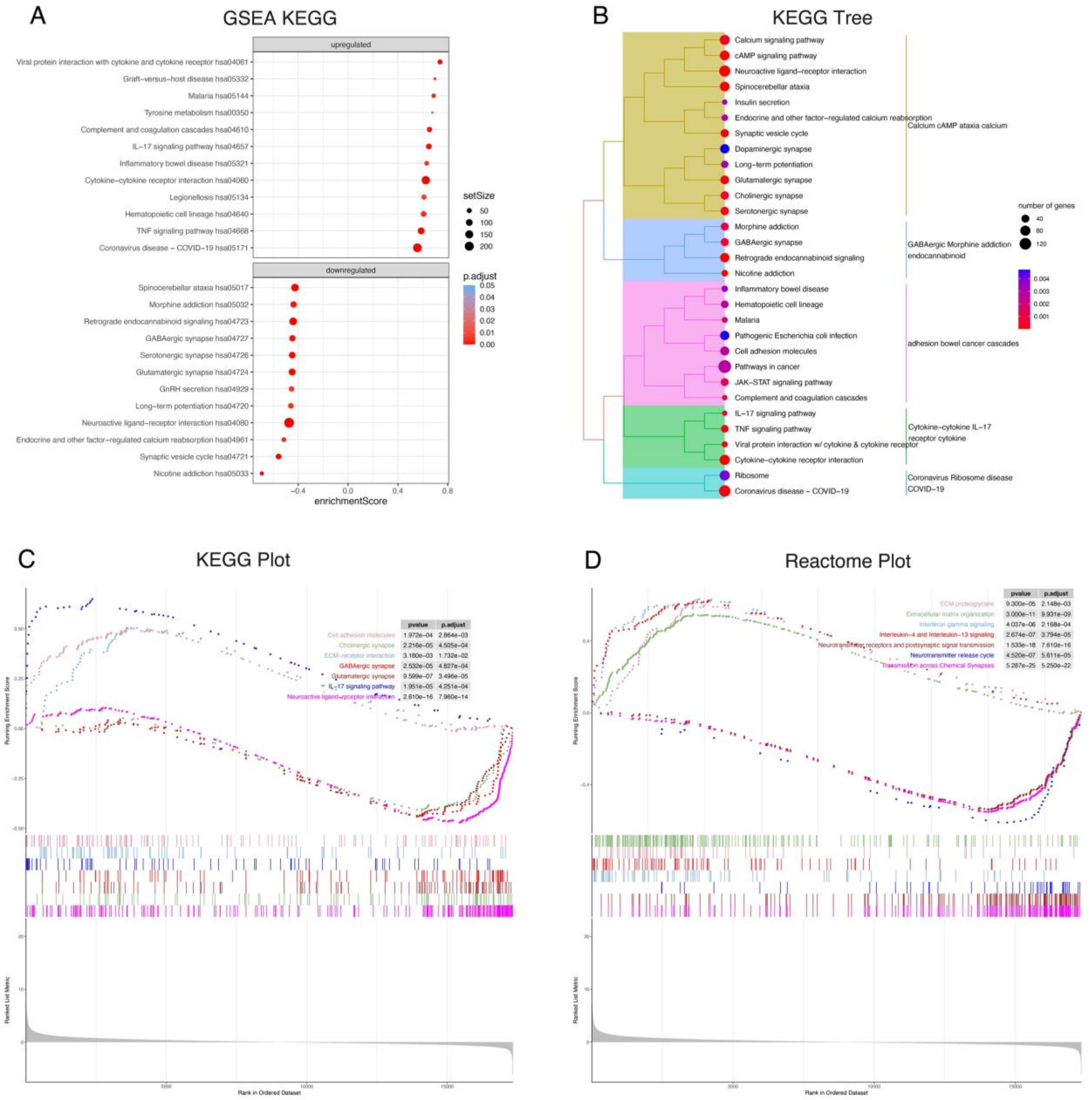


**Supplemental Figure 1: GSEA Analysis Plots.** (A) GSEA KEGG analysis of differential gene expression between subjects with ASD and control subjects. (B) GSEA KEGG Tree and KEGG Plot (C) of differentially expressed pathways in subjects with ASD. (D) Reactome Plot of differentially expressed pathways in subjects with ASD.

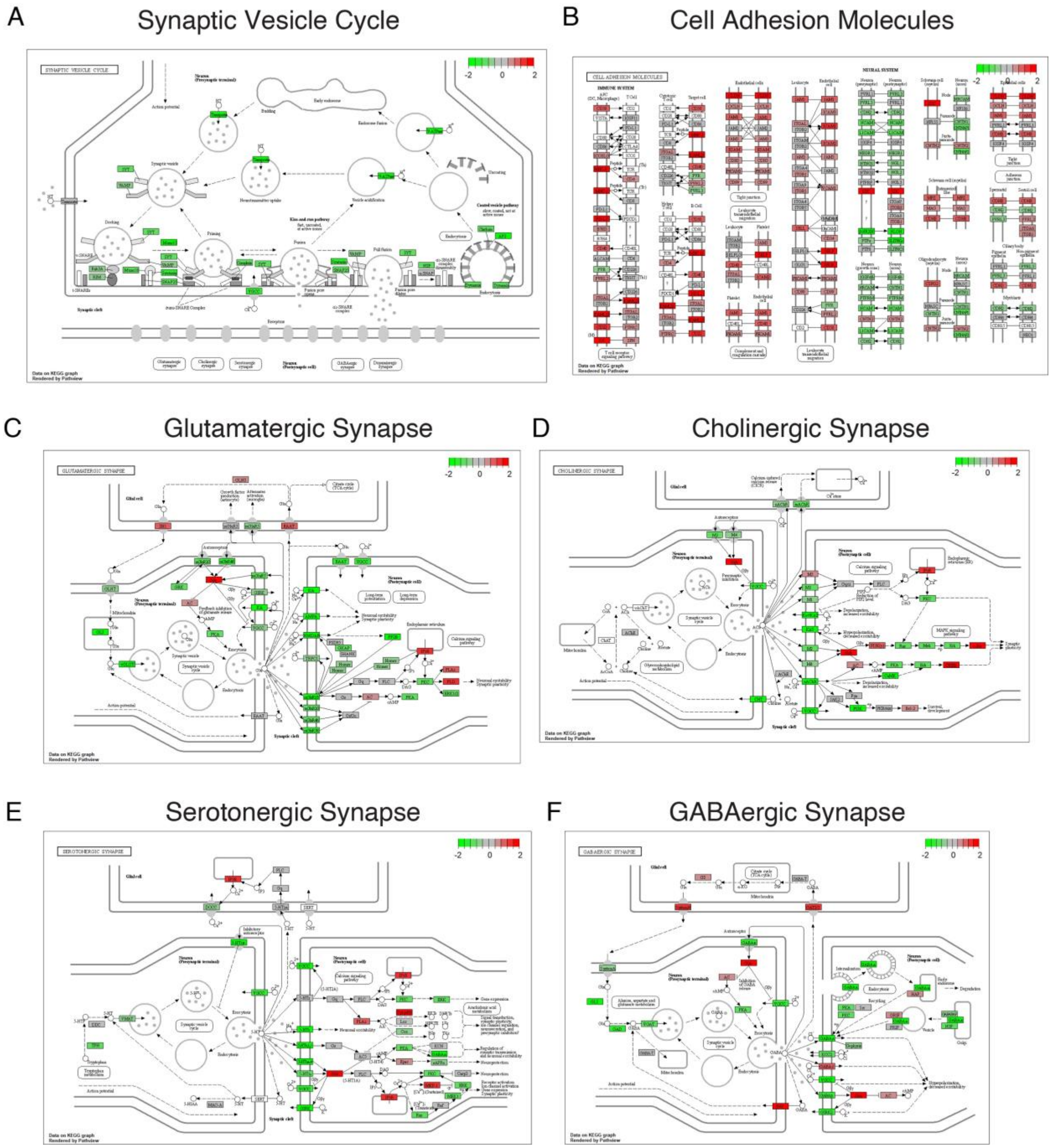


**Supplemental Figure 2: Synaptic Pathways Altered in the Hippocampus of Children with ASD.** RNAseq analysis identified differentially expressed genes in pathways involved in synaptic vesicle cycle (A), cell adhesion (B), glutamatergic synapses (C) cholinergic synapses (D) serotonergic synapses (E) and GABAergic synapses (F).

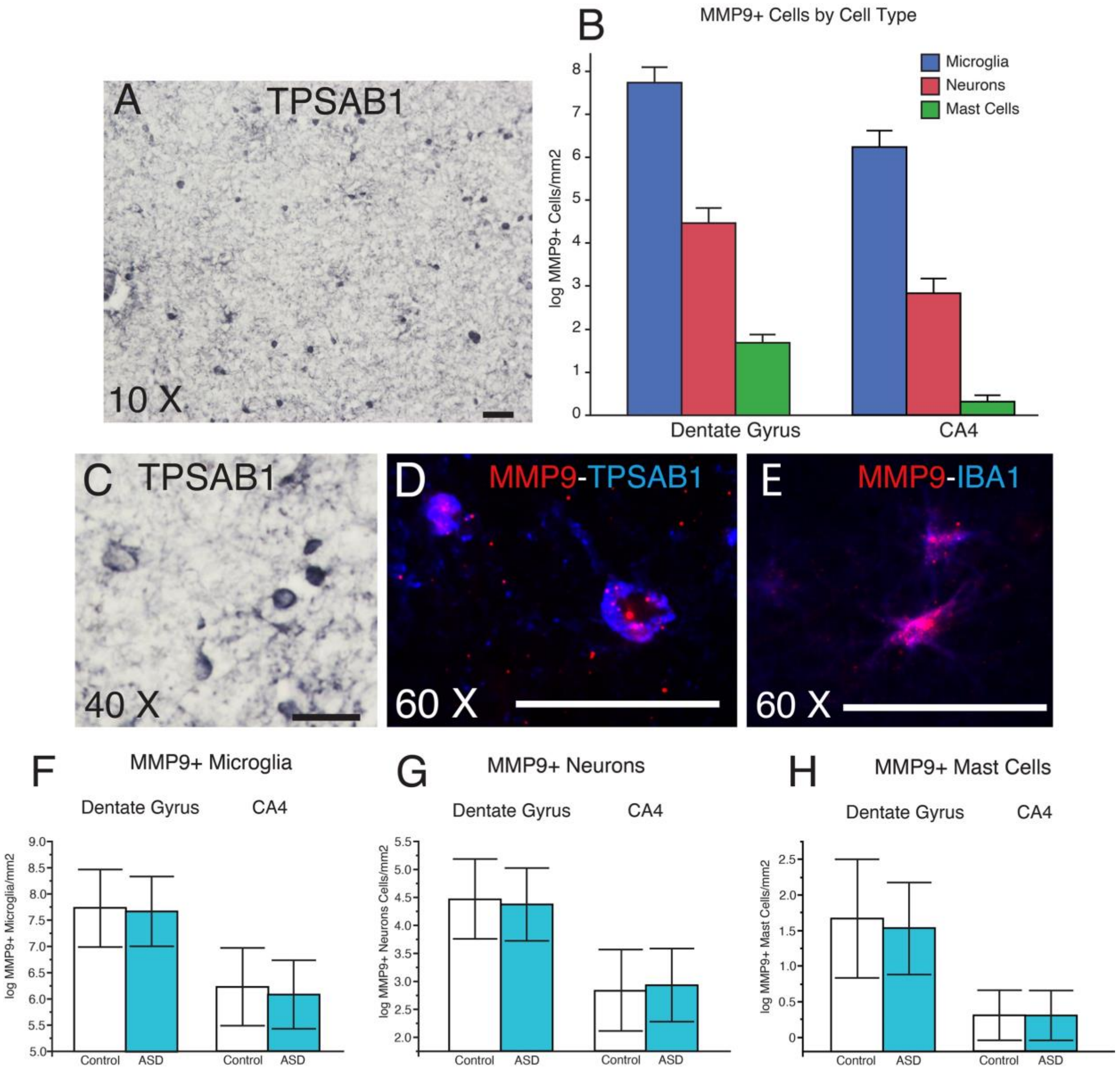


**Supplemental Figure 3: Cell type expression of MMP9 in the Human Hippocampus.** (A) Mast cells were detected in the human hippocampus of control subjects with the mast cell specific marker tryptase alpha/beta 1 (TPSAB1). Microscopy analysis of MMP9 labeling by cell type determined by morphology demonstrated that microglia are the predominant population of MMP9 labeled cells in the hippocampus, followed by neurons, and a small number of mast cells (B). (C) Representative micrograph of MMP9 labeled cells with a mast cell morphology in the human hippocampus. Dual immunofluorescence demonstating MMP9 expression in TPSAB1 mast cells (D) and IBA1 positive microglia (E). Scale bars = 50 μm. No differences were detected between subjects with ASD and non-ASD control subjects for densities of MMP labeled microglia, neurons, or mast cells (F-G). Significance values are derived from stepwise linear regression models. Bar graphs depict the mean (histogram) and 95% confidence intervals (black lines).

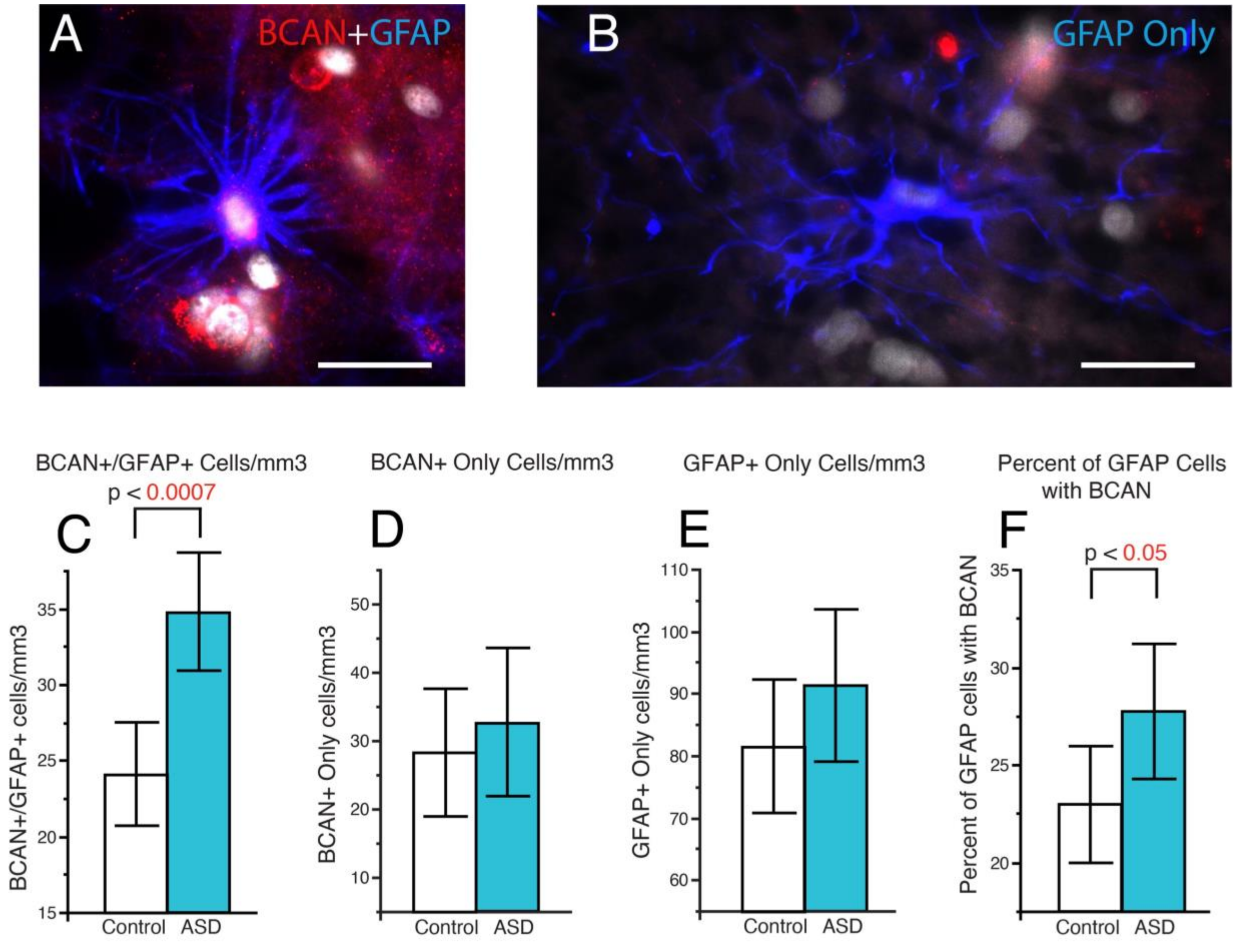


**Supplemental Figure 4: Increased Density of Astrocytes Co-labeled with Brevican in the Hippocampus of Children with ASD.** Dual immunofluorescence labeling was used to quantify the numerical density of GFAP positive astrocytes with or without brevican (BCAN) labeling in a subset of samples from children with ASD and non-ASD control subjects (n=6/group). Photomicrographs depicting an astrocyte labeled with GFAP (blue) and BCAN (red) (A) and an astrocyte labeled with GFAP only (B). The density of BCAN+/GFAP+ astrocytes was significantly increased in subjects with ASD compared to control subjects (C). In comparison, no difference was detected for astrocytes labeled with BCAN only (D) or GFAP only (E). Increased percentage of GFAP astrocytes were co-labeled with BCAN in subjects with ASD (F). Significance values are derived from stepwise linear regression models. Bar graphs depict the mean (histogram) and 95% confidence intervals (black lines).

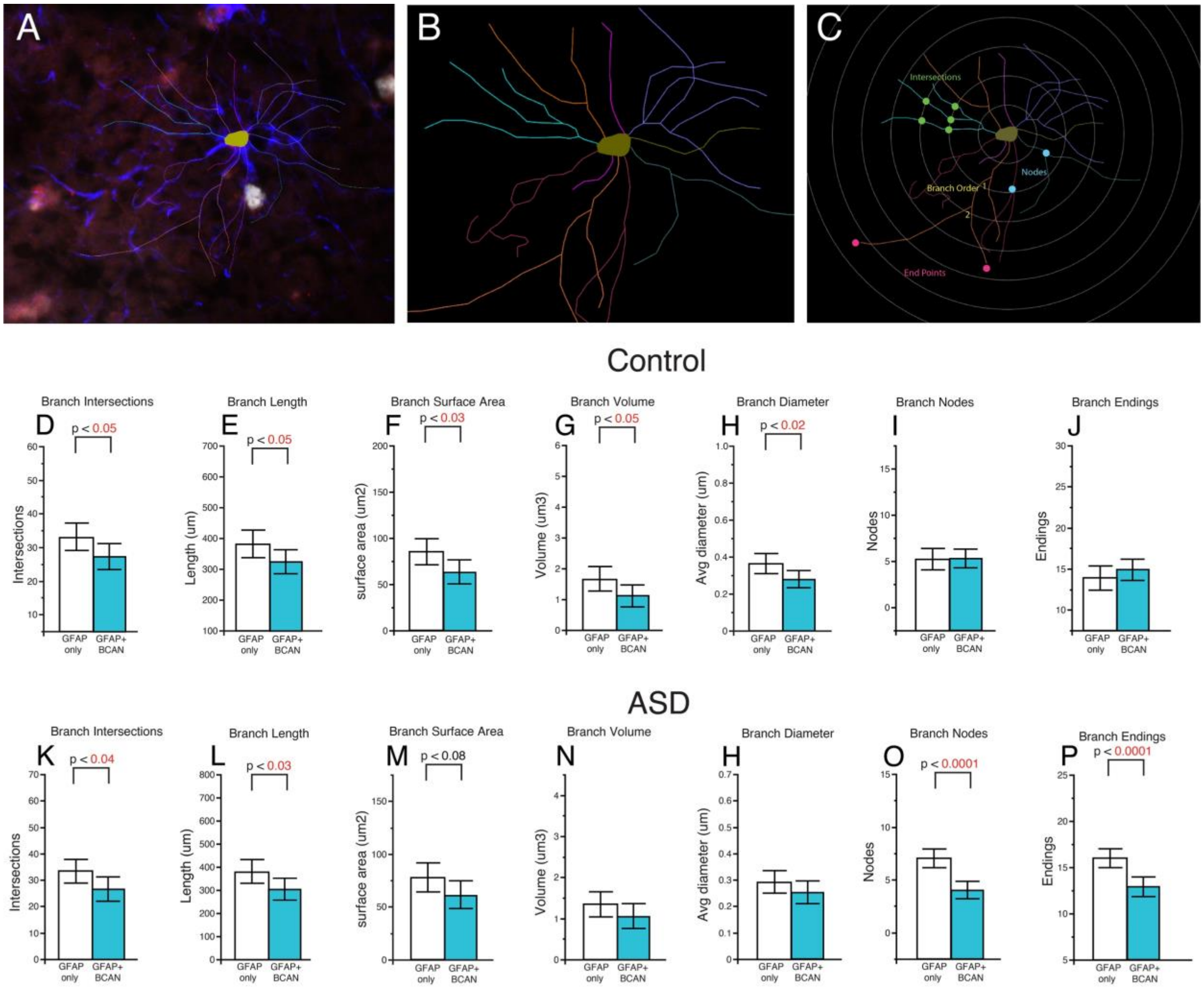


**Supplemental Figure 5: Brevican positive Astrocytes Display Immature Glial Cell Morphology.** Sholl analysis was conducted on a subset of samples from children with ASD and non-ASD control subjects (n=6/group). The cell body and branches of GFAP positive astrocytes were traced and analyzed using Sholl analysis with Neurolucida 360 software (A-C). Glial cells were also classified on the basis of co-expression of brevican (BCAN). Glial cells co-labeled with BCAN from control subjects had significant decreases in branch intersections, branch length, surface area, volume, and diameter (D-J). In comparison, glial cells co-labeled with BCAN from subjects with ASD had significant decreases in branch intersections, branch length, nodes, and endings (K-P). Significance values are derived from stepwise linear regression models. Bar graphs depict the mean (histogram) and 95% confidence intervals (black lines).
